# Serial intervals observed in SARS-CoV-2 B.1.617.2 variant cases

**DOI:** 10.1101/2021.06.04.21258205

**Authors:** Rachael Pung, Tze Minn Mak, CMMID COVID-19 working group, Adam J Kucharski, Vernon J. Lee

## Abstract

Rapid growth of the SARS-CoV-2 variant B.1.617.2 has been observed in many countries. The factors driving the recent rapid growth of COVID-19 cases could be attributed to shorten generation intervals or higher transmissibility (effective reproduction number, R), or both. Establishing the reasons for the observed rapid growth is key for outbreak control. In this study, we analysed the serial interval of household transmission pairs infected with SARS-CoV-2 B.1.617.2 variant and compared with those who were infected prior to the occurrence of the major global SARS-CoV-2 variants. After controlling for confounding factors, our findings suggest no significant changes in the serial intervals for SARS-CoV-2 cases infected with the B.1.617.2 variant. This, in turn, lends support for the hypothesis of a higher R in B.1.617.2 cases.

## Main

The SARS-CoV-2 lineage B.1.617 was declared a Variant of Concern by the World Health Organization given preliminary evidence suggesting faster spread relative to other circulating variants.^1^ However, the epidemiological factors contributing to this difference remain unclear. In particular, an increase in observed growth rate could be the result of a shorter generation interval (i.e. delay from one infection to the next) or an increase in the effective reproduction number, R, of an infected individual, or both.^2^ Whereas a shorter generation interval would increase the speed but not magnitude of individual-level transmission, a larger value of R would require a higher level of vaccination or physical distancing to suppress transmission.

In Singapore, whole genome sequencing is performed for respiratory samples from COVID-19 cases who tested positive by PCR with a cycle threshold of 30 and below. The B.1.617.2 variant was first identified in local cases on Apr 27, 2021. Despite high levels of adherence to mask wearing and physical distancing in the country,^3,4^ clusters of B.1.617.2 were detected and some clusters displayed rapid growth of infections within a short period of time.

We investigated possible drivers of B.1.617.2 growth by studying the serial intervals (i.e. onset-to-onset delay) — a proxy for the generation interval — between pairs of a primary case and a secondary case occurring among household members. Exposure histories were reviewed for all household transmission pairs involving COVID-19 cases infected with the B.1.617.2 variant or cases identified between Apr 27 to May 22, 2021 and whose samples were yet to be or could not be sequenced. The B.1.617.2 variant was detected in 97% of the sequenced samples from local COVID-19 cases identified in this period. Secondary cases with potential exposure to (i) more than one primary case in the household or (ii) to other cases outside the household were omitted from analysis. Households with secondary cases having different symptom onset dates were also omitted from the analysis as we were unable to rule out multiple generations of transmission.

For comparison, we identified household transmission pairs prior to the partial lockdown in Singapore on Apr 7, 2020 and applied the same exclusion criteria. This time period precedes the occurrence of the major global SARS-CoV-2 variants^5^ and most closely matches the social activity and workplace arrangements in Apr 2021.^6^ For a given time from symptom onset to isolation of a recent primary case, we randomly sampled primary cases with the same time from onset to isolation prior to the partial lockdown in Apr 2020 and sampled the corresponding serial intervals. We fitted a skewed normal distribution to the set of serial intervals to account for negative serial intervals arising from pre-symptomatic transmission and the process was repeated 1000 times to obtain a mean and 95% confidence interval. The result is a distribution of serial intervals observed in cases prior to the occurrence of SARS-CoV-2 variants while controlling for the same duration from symptoms onset to isolation of the primary case as observed in the recent primary cases.

32 recent household transmission pairs were identified, of which 28 were associated with the B.1.617.2 variant. The median duration from symptoms onset to isolation of the recently notified primary cases was three days (IQR 2–4) (Figure 1a). This was 3.0 days (95% CI 2–4) in the 63 household transmission pairs identified before Apr 7, 2020 after adjustment (Figure 1b and c). The mode of the serial interval was two days for B.1.617.2 cases and for all the recently notified cases and 2.8 days (95% CI -1–4) for cases prior to the lockdown.

**Figure 1.**
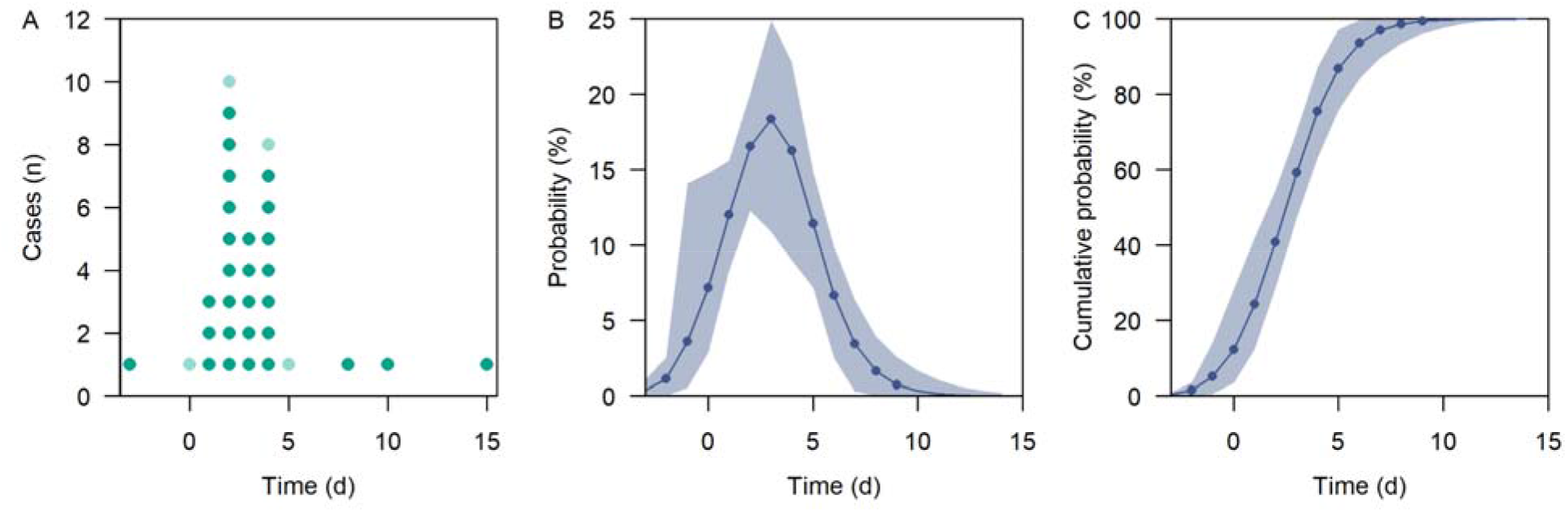
Serial interval of household transmission pairs. (a) B.1.617.2 cases (green) or cases identified between Apr 27 to May 22, 2021 and whose samples were yet to be sequenced (light green), (b) probability density function of serial interval of cases identified prior to the partial lockdown in Apr 7, 2020, (c) cumulative density function (b). Majority of the primary cases had known exposure(s) outside the household and secondary cases do not have the same exposure as the primary case thereby allowing us to identify the directionality of infection. Negative serial intervals, which signifies pre-symptomatic transmission are also included in the analysis.

Our preliminary investigations of the recent, albeit small dataset of, B.1.617.2 cases, suggest no significant changes in serial intervals. In turn, this lends support to the hypothesis that the recent rapid growth is potentially driven by an increase in the average number of secondary cases generated by a case infected with the B.1.617.2 variant. Studies with proper control of confounding factors are thus crucial to tease out the key epidemiological factors that facilitate the increased transmissibility of the B.1.617.2 variant. Without signs of lowered disease severity for B.1.617.2, contact tracing and testing around COVID-19 cases, along with vaccination and non-pharmaceutical interventions, continue to remain key SARS-CoV-2 outbreak control measures in the short term.

## Data Availability

All code and data to reproduce the analysis can be found at https://github.com/rachaelpung/serial_interval_covid_b.1.617.2

## Code and data

https://github.com/rachaelpung/serial_interval_covid_b.1.617.2

