## Supplementary material for "Serial intervals observed in SARS-CoV-2 B.1.617.2 variant cases": Figure 1

[1] Ministry of Health, Singapore
[2] Centre for the Mathematical Modelling of infectious Diseases, and Department of Infectious Disease Epidemiology, London School of Hygiene and Tropical Medicine, London
[3] National Public Health Laboratory, National Centre for Infectious Diseases, Singapore
[4] Saw Swee Hock School of Public Health, National University of Singapore, Singapore
*Listed at the end of supplementary


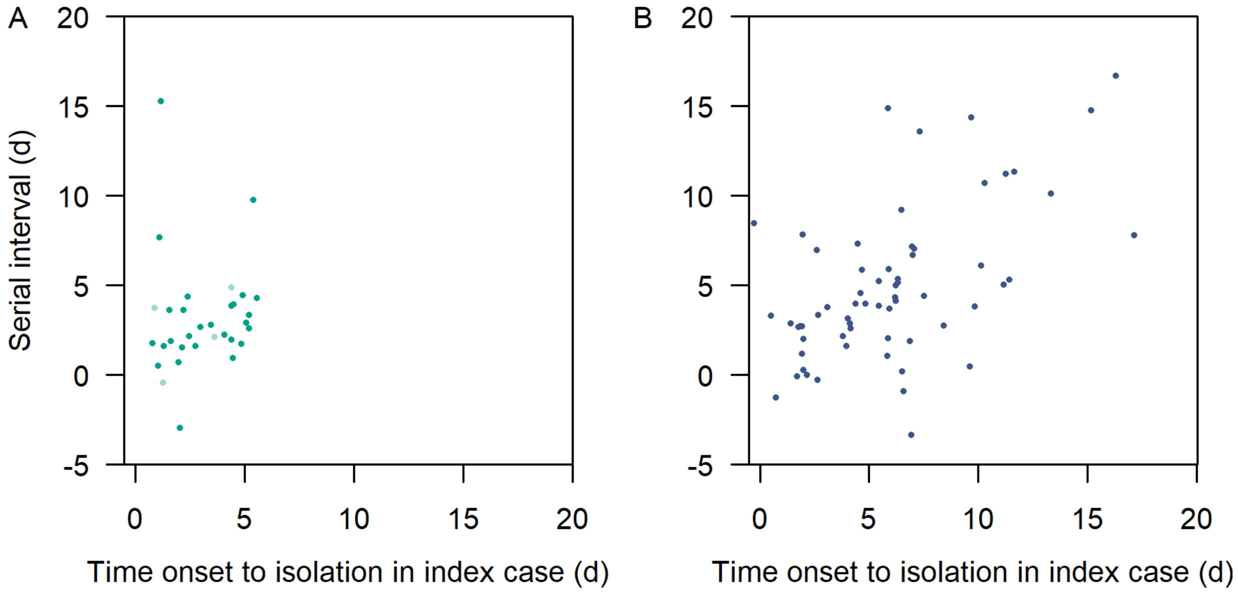


Supplementary figure 1 Time from onset to isolation in primary case against the serial interval in household pairs (a) in recent B.1.617.2 cases (green) and cases identified between Apr 27 to May 22, 2021 and whose samples were yet to be sequenced (light green), (b) in cases identified prior to the partial lockdown in Apr 7, 2020.

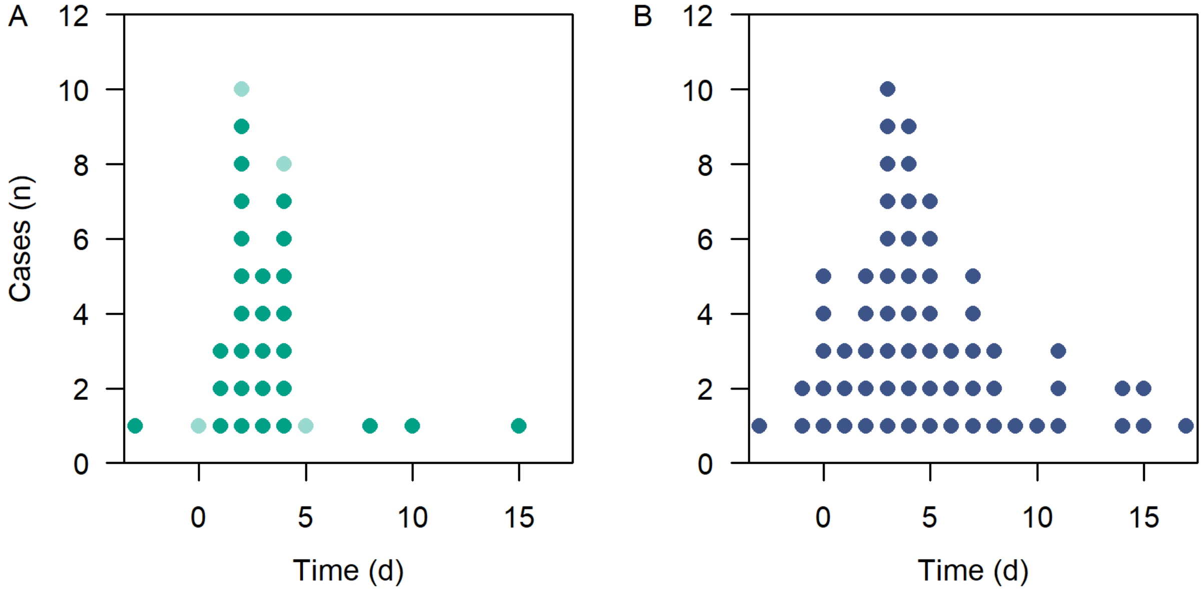


Supplementary figure 2 Serial interval of household transmission pairs. (a) B.1.617.2 cases (green) or cases identified between Apr 27 to May 22, 2021 and whose samples were yet to be sequenced (light green), (b) in cases identified prior to the partial lockdown in Apr 7, 2020 without adjusting for time from onset to isolation in primary case.

**CMMID COVID-19 working group**

The following authors were part of the Centre for Mathematical Modelling of Infectious Disease COVID-19 Working Group. Each contributed in processing, cleaning and interpretation of data, interpreted findings, contributed to the manuscript, and approved the work for publication: Kathleen O'Reilly, Gwenan M Knight, Lloyd A C Chapman, Sam Abbott, Carl A B Pearson, James D Munday, Yalda Jafari, Yang Liu, Rachel Lowe, Hamish P Gibbs, Simon R Procter, Sebastian Funk, Nikos I Bosse, Graham Medley, C Julian Villabona-Arenas, Nicholas G. Davies, Kaja Abbas, Alicia Rosello, Christopher I Jarvis, Stefan Flasche, Amy Gimma, Rosalind M Eggo, Oliver Brady, Stéphane Hué, Billy J Quilty, Damien C Tully, W John Edmunds, Samuel Clifford, Katherine E. Atkins, Mark Jit, Anna M Foss, Sophie R Meakin, Ciara V McCarthy, Paul Mee, Frank G Sandmann, William Waites, Mihaly Koltai, Kiesha Prem, Joel Hellewell, Emilie Finch, Timothy W Russell, Matthew Quaife, Katharine Sherratt, Fiona Yueqian Sun, Rosanna C Barnard, Kerry LM Wong, Akira Endo, David Hodgson.

The following funding sources are acknowledged as providing funding for the working group authors. This research was partly funded by the Bill & Melinda Gates Foundation (INV-001754: MQ; INV-003174: KP, MJ, YL; INV-016832: SRP; NTD Modelling Consortium OPP1184344: CABP, GFM; OPP1139859: BJQ; OPP1191821: KO'R). BMGF (INV-016832; OPP1157270: KA). CADDE MR/S0195/1 & FAPESP 18/14389-0 (PM). EDCTP2 (RIA2020EF-2983-CSIGN: HPG). ERC Starting Grant (#757699: MQ). ERC (SG 757688: CJVA, KEA). This project has received funding from the European Union's Horizon 2020 research and innovation programme - project EpiPose (101003688: AG, KLM, KP, MJ, RCB, WJE, YL). FCDO/Wellcome Trust (Epidemic Preparedness Coronavirus research programme 221303/Z/20/Z: CABP). This research was partly funded by the Global

Challenges Research Fund (GCRF) project 'RECAP' managed through RCUK and ESRC (ES/P010873/1: CIJ). HDR UK (MR/S003975/1: RME). HPRU (This research was partly funded by the National Institute for Health Research (NIHR) using UK aid from the UK Government to support global health research. The views expressed in this publication are those of the author(s) and not necessarily those of the NIHR or the UK Department of Health and Social Care200908: NIB). MRC (MR/N013638/1: EF; MR/V027956/1: WW). Nakajima Foundation (AE). NIHR (16/136/46: BJQ; 16/137/109: BJQ, FYS, MJ, YL; 1R01AI141534-01A1: DH; NIHR200908: LACC, RME; NIHR200929: CVM, FGS, MJ, NGD; PR-OD-1017-20002: AR, WJE). Royal Society (Dorothy Hodgkin Fellowship: RL). UK DHSC/UK Aid/NIHR (PR-OD-1017-20001: HPG). UK MRC (MC_PC_19065 - Covid 19: Understanding the dynamics and drivers of the COVID-19 epidemic using real-time outbreak analytics: NGD, RME, SC, WJE, YL; MR/P014658/1: GMK). UKRI (MR/V028456/1: YJ). Wellcome Trust (206250/Z/17/Z: TWR; 206471/Z/17/Z: OJB; 208812/Z/17/Z: SC, SFlasche; 210758/Z/18/Z: JDM, JH, KS, SA, SFunk, SRM; 221303/Z/20/Z: MK). No funding (AMF, DCT, SH).
